# SPIRIT-CONSORT-TM: a corpus for assessing transparency of clinical trial protocol and results publications

**DOI:** 10.1101/2025.01.14.25320543

**Authors:** Lan Jiang, Colby J Vorland, Xiangji Ying, Andrew W Brown, Joe D Menke, Gibong Hong, Mengfei Lan, Evan Mayo-Wilson, Halil Kilicoglu

## Abstract

Randomized controlled trials (RCTs) can produce valid estimates of the benefits and harms of therapeutic interventions. However, incomplete reporting can undermine the validity of their conclusions. Reporting guidelines, such as SPIRIT for protocols and CONSORT for results, have been developed to improve transparency in RCT publications. In this study, we report a corpus of 200 RCT publications, named SPIRIT-CONSORT-TM, annotated for transparency. We used a comprehensive data model that includes 83 items from SPIRIT and CONSORT checklists for annotation. Inter-annotator agreement was calculated for 30 pairs. The dataset includes 26,613 sentences annotated with checklist items and 4,231 terms. We also trained natural language processing (NLP) models that automatically identify these items in publications. The sentence classification model achieved 0.742 micro-F1 score (0.865 at the article level). The term extraction model yielded 0.545 and 0.663 micro-F1 score in strict and lenient evaluation, respectively. The corpus serves as a benchmark to train models that assist stakeholders of clinical research in maintaining high reporting standards and synthesizing information on study rigor and conduct.

## Background & Summary

Randomized controlled trials (RCTs) are foundational to evidence-based medicine^1^. When well-designed and rigorously conducted, RCTs can provide valid estimates of effects of therapeutic interventions^2^. For RCTs to benefit clinical practice and health policy, they must be reported thoroughly and transparently^2,3^. Complete reporting facilitates the assessment of RCT validity and applicability^2^. Given the high cost and time investment of RCTs, transparent reporting also helps avoid unnecessary duplication and research waste^2,4^. Unfortunately, even well-conducted RCTs often suffer from inadequate reporting^2,5,6^.

The SPIRIT 2013 Statement^7,8^ and CONSORT 2010 Statement^2,9^ are reporting guidelines that aim to enhance the reporting quality of RCT protocols and results, respectively. CONSORT 2010 (referred to as CONSORT for brevity, henceforth) consists of a checklist and a participant flowchart. The checklist includes 25 items essential for understanding the design, implementation, analysis, and results of parallel RCTs. CONSORT has been widely endorsed by journals, publishers, and editorial organizations, and its adoption has been found to be positively correlated with completeness of reporting^10,11^. However, studies have also repeatedly shown that key methodological details like allocation concealment remain poorly reported even in articles published in endorsing journals^5,6^. An overview of systematic reviews found that CONSORT adherence was reported to be inadequate in 88% of the reviews^6^. The complementary SPIRIT 2013 guidelines^7,8^ (referred to as SPIRIT, henceforth) consist of recommended items and a figure to be included in trial protocols. SPIRIT includes many applicable items from CONSORT, especially items related to methodology, and often encourages authors to report more information than would typically be included in a results report. Protocols are widely used to appraise trial conduct by funding agencies, institutional review board, regulatory agencies, and systematic reviewers^7,8^. Ensuring that a trial protocol is rigorous and transparent before the trial begins can improve the execution of the trial and minimize protocol amendments, ultimately translating into more reliable trial results. Comparison of protocols with results publications can also pinpoint issues in trial conduct, such as outcome switching^12^.

Low adherence to CONSORT and SPIRIT demonstrates that journal endorsement does not guarantee that authors will report the minimum recommended information. Manually verifying that the authors have adhered to CONSORT recommendations has been shown to improve reporting^10,11^ but is not scalable beyond a small number of well-resourced journals. Automatic screening for SPIRIT and CONSORT compliance could allow more journals to assess reporting quality, reduce burden on editors and peer reviewers, and enhance RCT reporting quality. Natural language processing (NLP) and machine learning (ML) techniques can support such automatic screening tools^13–16^.

There has been significant NLP research targeting RCT publications, primarily for use in systematic reviews and evidence synthesis^17^. This includes classifying sentences in abstracts or full-text articles by PICO elements (Population, Intervention, Comparator, and Outcome) for article screening^18–21^, extracting PICO-related or other methodological terms to aid data extraction^22–28^, and classifying text for automated risk-of-bias assessment^29–31^. These studies often focus on a small number of elements relevant to trials and they do not specifically consider reporting quality. Other studies focus on annotating and extracting clinical trial data from registries; for example, the Chia corpus provides fine-grained annotations of eligibility criteria from ClinicalTrials.gov^32^. NLP work focusing specifically on RCT reporting transparency is relatively recent. In prior work, we constructed CONSORT-TM, a corpus of 50 RCT results publications annotated for CONSORT checklist items at fine granularity (37 items)^14^. We also trained and validated NLP models based on this corpus which label individual sentences for the checklist items they report^14,33,34^. Additionally, we applied a model that specifically focuses on methodology-related CONSORT items at large scale (176,469 publications) to study RCT reporting patterns over time, which showed that methodology reporting in RCT publications had improved over time but that it remained suboptimal for many items^35^.

Although CONSORT-TM and the models trained on it enable automated screening of RCT publications, they have several shortcomings. First, the corpus is relatively small (5,246 annotations over 4,845 sentences). Second, the best NLP model currently yields 0.71 micro-F1 and 0.67 macro-F1 at the sentence level and fails on some infrequent labels partly due to small training size. This limits the practical applicability of the models. Third, adherence to CONSORT can have limited effect on improving the rigor and conduct of a trial, because by the time the results are reported in a manuscript, it may be too late to improve the trial design and conduct.

In this work, we aim to address some of these limitations by expert annotation of a larger corpus that not only focuses on results publications but also protocols of clinical trials and includes a larger and more granular set of checklist items than considered before. Our combined annotation scheme recognizes the overlap between SPIRIT and CONSORT. A major motivation for our expansion is the recent proposal to better align CONSORT and SPIRIT to enhance usability, implementation, and efficiency^36^, which our corpus and models also support. In sum, our contributions are as follows:

- We have designed a comprehensive data model of RCT reporting characteristics based on SPIRIT and CONSORT guidelines (83 items).
- We have annotated the largest corpus of RCT protocol-results publication pairs, to our knowledge, using the data model and made it publicly available from https://github.com/ScienceNLP-Lab/RCT-Transparency/tree/main/SPIRIT-CONSORT-TM.
- We trained and validated strong baseline models based on state-of-the-art neural network architectures for article, sentence, and term level recognition of RCT characteristics.

The corpus can serve as a benchmark to support further development of NLP models that support automated transparency screening of RCT publications.

## Methods

### Trial selection

Our search and screening steps are visualized in Figure 1. We included parallel group RCTs of interventions because CONSORT applies to parallel group trials. We excluded pilot and feasibility studies for which other reporting guidelines are available. To be included in the study, trials must have been registered on ClinicalTrials.gov and must have published both a study protocol and a manuscript reporting the primary results. We used stratified random sampling to identify eligible protocols, as previously described^37^. On August 10, 2022, we searched for trial protocols from January 2011 to August 2022 on PubMed Central. Detailed inclusion and exclusion criteria, along with the search strategy, are located at https://osf.io/8rg4h/.

**Figure 1.**
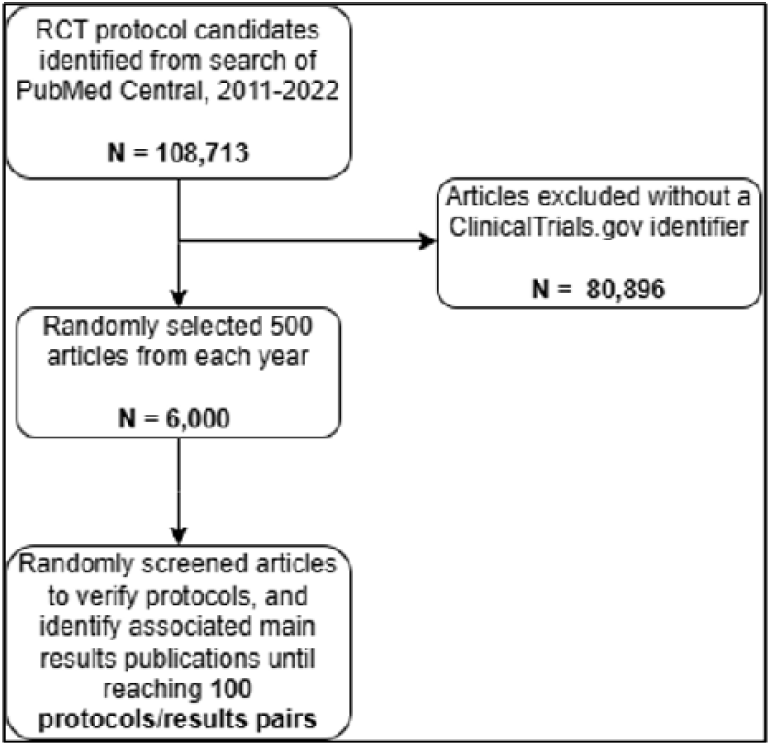
Flow chart of searching and screening process.

We retained articles with a ClinicalTrials.gov identifier in the abstract or full text (excluding references) using regular expression pattern matching. We then randomly selected 500 articles from each year, yielding 6000 citations. After randomly shuffling the order, we screened citations in duplicate and resolved discrepancies through discussion. For each included protocol, we identified the earliest main results publication by reviewing linked publications on ClinicalTrials.gov and applying the eligibility criteria. We continued screening records until we reached 100 included protocol/results pairs (100 protocols and 100 main results publications). Our search concluded in September 2022.

### Data annotation and curation

Based on the SPIRIT^7,8^ and CONSORT^2,9^ guidelines, we developed an annotation guide, also available at https://osf.io/8rg4h/. We operationalized a total of 83 items from both guidelines, including four applicable to protocols only, eleven applicable to results only, and 78 applicable to both. We assigned a number and short description to each item (e.g., *11a_Intervention_Description*). We developed guidance for annotating for each item, along with examples. We updated the annotation guide throughout the annotation process to reflect protocol changes and to refine instructions. Several items from each checklist were excluded from annotation. These items and the rationale for exclusion are provided below:

- SPIRIT: 2b (Information from the World Health Organization Trial Registration Data Set) and 3 (Protocol version) are almost never reported in published protocols. 6a and 6b (Background and rationale) are broad and subjective, so we did not believe they could be assessed reliably.
- CONSORT: 2a (Background), 20 (Limitations) and 22 (Interpretation) are also broad and subjective.

We downloaded protocol and results publications for 100 trials from PubMed Central as HTML files and converted them to plain text for annotation. We completed the annotations using the brat annotation tool (version 1.3)^38^, which allows span-based text annotations. Because brat does not preserve article structure, hashtags were used to indicate section headers and their depth (e.g., # for top level headers, ## for headers of their subsections).

After span-based annotation of checklist items in brat, we constructed the final corpus by automatically converting span annotations to article-level, sentence-level, and term-level datasets. The article-level dataset simply includes information on whether a checklist item is reported in an article (binary labels). All 83 items are included in this dataset. The sentence-level dataset is multi-label and includes individual sentences associated with one or more checklist items (or none). All items except *2_Abstract_structured* (whether the publication includes a structured abstract) are included in the sentence-level dataset. The term-level dataset includes word/phrase span annotations that precisely describe the checklist items. 22 items are considered for this dataset. Each dataset could serve a different purpose. Specifically, the article-level dataset is appropriate for developing text classification models to assess whether checklist items are reported in an article or not, while sentence-level dataset allows development of models that identify relevant sentences for each item as well. On the other hand, the term-level dataset is appropriate for developing information extraction models that identify specific RCT characteristics (e.g., sequence generation method) that can help describe the trial conduct (e.g., to assess risk of bias). We consider the sentence-level dataset as the primary dataset in our corpus.

To facilitate efficient annotation in brat, we distinguished three options for annotating spans:

- *Section annotation*: a section header span relevant to a checklist item is annotated, to indicate that all sentences within that section are relevant to the item (e.g., The section header “Primary outcomes” is annotated to indicate the label *12a_Outcome_Definitions* for all sentences in that section.)
- *Trigger annotation*: a word/phrase relevant to an item is annotated, to indicate that the enclosing sentence contains information related to the item (e.g., the span “The specific aim” is annotated to indicate that the enclosing sentence relates to the item *7_Objectives*.)
- *Term annotation*: a word/phrase that precisely describes the item is annotated (e.g., “NCT01243554” to indicate the item *3a_Registry_Number*.)

During brat annotation, we used the suffix *_Term* in item labels (e.g., *3a_Registry_Number_Term*) to indicate that the annotator should annotate the item as a term. All other items could be annotated as section or trigger spans. Section annotation helps speed up the annotation process and reduces annotator burden, because instead of labeling every sentence in a relevant section, only the section header is annotated. This is particularly useful for commonly reported items that have multiple pieces and often reported in specific sections (e.g., *11a_Intervention_Description, 12a_Outcome_Definitions*). All 83 items are described in the annotation guide.

We converted brat span annotations to final article-level and sentence-level datasets using an automated label propagation process. No specific post-processing is needed for the term-level dataset. In the article-level dataset, the items that were annotated in an article were recorded as 1 (present) and those that were not as 0 (absent). For the sentence-level dataset, section header annotations were propagated down to all sentences in that section, unless the sentence was annotated with a different label. There are 11 exceptions to this rule tied to specific labels determined by the annotators and listed in the annotation guide. For example, if the sentence is labeled with *18a_Data_Collection*, and the section header with *12a_Outcomes_Definitions*, the sentence is still additionally labeled with *12a_Outcomes_Definitions*. Labels annotated as triggers and terms were applied to the enclosing sentences. We included table contents (i.e., rows) in brat annotation; however, because there were not many row annotations and rows are often quite different from natural language sentences, we simply associated the labels on table rows with the table captions, which are treated as regular sentences, and excluded table rows from the sentence-level and term-level datasets. Figure captions are also included in the corpus. Example brat annotations corresponding to different annotation options are shown in Figure 2.

**Figure 2.**
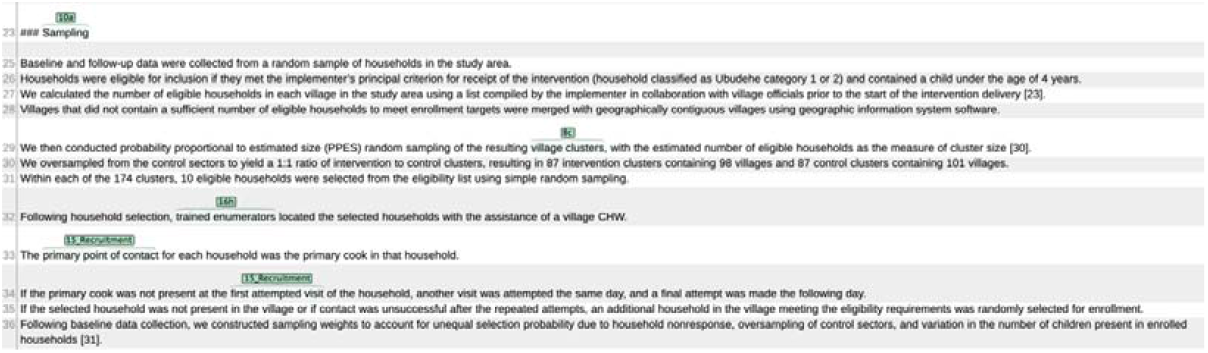
Brat annotation example (from the publication with PubMed Central identifier PMC6546207) illustrating section, trigger, and term level annotations. *10a_Participants_Inclusion* is annotated by the section header span “Sampling”. This indicates a section-level annotation and, as a result, all the shown sentences are automatically annotated as *10a_Participants_Inclusion* through label propagation in post-processing. *15_Recruitment* and *16h_Personnel_Enrollment* are annotated at the trigger level; therefore, sentence 32 gets the label *16h_Personnel Recruitment* and sentence 33 and 34 the label *15_Recruitment* in post-processing. *8c_Design_Centers* is a term-level item; so, sentence 29 gets this label in sentence-level dataset and the phrase “village clusters” gets the label in term-level dataset. Note that because of label propagation in post-processing, some sentences will have multiple labels, e.g., sentence 29 gets both labels *10a_Participants_Inclusion* and *8c_Design_Centers*. All four items are labeled as 1 in the article-level dataset. Some labels are abbreviated by brat to improve readability.

Annotation was performed by four experts (CJV, XY, AWB, and EM-W) in epidemiology and meta-science (the study of the scientific process), with expertise in evaluating the quality and transparency of research studies and specific experience in assessing RCT protocols and results publications. Annotators were always assigned to pairs of protocols and results publications for within-trial consistency.

For the first 30 pairs of articles, annotators worked in duplicate, and any disagreements were resolved through discussion. We also calculated inter-annotator agreement for these articles. Three annotators annotated the first five pairs (001-005) of articles (CJV, AWB, and EM-W). During this phase, the annotators had discussions to resolve discrepancies and to develop and finalize the annotation guide. For the next 14 pairs of articles (006-019), each pair of articles was first annotated individually by two (of three) annotators and then reconciled by the annotators. Subsequently, four annotators were involved in annotation of the next 11 pairs of articles (020-030). The fourth annotator (XY) reconciled the first five pairs of articles after being paired with other annotators for reconciliation. Double annotation ensured consistency between annotators. After this iteration, each pair of articles in the remaining 70 pairs was annotated by a single annotator. We considered the reconciled version to construct the ground truth datasets for modeling.

### Data Records

The annotated SPIRIT-CONSORT-TM corpus is available from https://github.com/ScienceNLP-Lab/RCT-Transparency/tree/main/SPIRIT-CONSORT-TM, including a *README*.*md* file that explains the corpus organization. To build the corpus, we converted the raw brat annotation data to article-level, sentence-level, and term-level datasets, as explained above. Raw brat data includes the label (checklist item), the annotated string, and its start and end character positions for each annotation.

The final corpus is organized as follows:

- *documents:* a directory that contains all 200 articles in plain text format. Files are organized under *Protocols* and *Results* subdirectories. The file names have the format *PairID_PMCID*.*txt*, where *PairID* is the pair number of the articles (001 to 100) and *PMCID* is the PubMed Central identifier of the article.
- *articles*.*csv*: a CSV (comma-separated values) file that includes the article-level dataset. Each row includes the following columns: *Protocol/Results* (whether the article is a protocol or results publication), *PairID, PMCID, ChecklistItem* (the checklist item identifier), *Reported* (1 if the article includes information related to the checklist item, 0 otherwise), and *Split* (train/valid/test).
- *sentences*.*csv*: a CSV file that includes the sentence-level dataset. In addition to *Protocol/Results, PairID, PMCID*, and *Split* columns, this file includes the following columns: *SentenceID* (sentence index within the article), *Sentence* (sentence text), *SentenceNoMarkers* (sentence text excluding header or table markers (#), *ChecklistItems* (a list that indicates the checklist item labels associated with the sentence), *SectionHeaders* (a list that includes all section headers associated with the sentence, from top level down to the innermost header), *IsSectionHeader* (whether the sentence is a section header itself; 0 or 1), *SentenceStartOffset* (the start position of the sentence in the article), and *SentenceEndOffset* (the end position).
- *terms*: This is a directory that includes term-level annotations, organized in two sub-directories: *raw_data* and *processed_data. raw_data* contains annotations in brat standoff annotation format. *processed_data* organizes the data into three JSONL files corresponding to training, validation, and test splits. For JSONL files, we follow the format for named entities in SciERC dataset^38^. Each row in a JSONL file corresponds to a single article that includes the following keys: *doc_key* (PMID/PMCID for each article), *sentences* (a list of tokens for each sentence), *ner* (a list of terms in the article, including their token-level start and end offsets and the corresponding checklist item labels), and *section_headers* (all section headers for each sentence).

Training, validation, and test sets were selected randomly (70 pairs for training, 10 pairs for validation, and 20 for test).

### Descriptive Statistics

High-level descriptive statistics of the corpus are provided in Table 1.

**Table 1.** High-level descriptive statistics of SPIRIT-CONSORT-TM. Mean and median values are per report. Std: standard deviation; IQR: inter-quartile range.

|  | Total No. | Mean (Std) | Median (IQR) |
| --- | --- | --- | --- |
| *Article-level* |  |  |  |
| Articles (protocols/results) | 200 (100/100) | - |  |
| Checklist items included | 8,285 | 41.43 (7.54) | 42.00 (10.25) |
| SPIRIT items in protocols | 2,841 | 28.41 (5.12) | 28.00 (7.00) |
| CONSORT items in results | 2,493 | 24.93 (3.27) | 25.00 (4.00) |
| Tokens | 1,211,107 | 6,055.54<br>(1,872.87) | 5,893.50<br>(1,946.50) |
| *Sentence-level* |  |  |  |
| Sentences | 52,294 | 261.47 (76.28) | 249.00 (94.50) |
| Annotated sentences | 26,613 | 133.07 (50.47) | 128.00 (57.75) |
| Annotations | 30,960 | 154.80 (58.74) | 147.00 (61.75) |
| *Term-level* |  |  |  |
| Annotations | 4,231 | 21.16 (7.37) | 20.50 (10.00) |
| Unique term mentions | 3,926 | 19.63 (6.49) | 20.00 (9.00) |
| Unique term types | 2,654 | 13.27 (3.05) | 14.00 (4.00) |

### Article-level dataset

We annotated a total of 200 articles (100 protocol-results pairs). The article-level dataset included 8,285 positive and 8,315 negative labels. On average, each article reports 41.43 (±7.54) items (out of 83), indicating that approximately half of all relevant items were reported in each article. On average, 61.8% of SPIRIT items considered were present in the protocol papers, while 75.6% of CONSORT items considered were reported in the results papers. This shows that results articles tend to report more comprehensively. CONSORT reporting in this corpus is similar to that observed in our previous study^14^, which showed that 74.3% of the items were reported. Item-specific statistics for the article-level dataset is provided in Supplementary Table S2. The most commonly annotated items included in both SPIRIT and CONSORT guidelines were eligibility criteria (*10a_Participant_Inclusion*), interventions (*11a_Intervention_Description*), outcome definitions (*12a_Outcome_Definitions*), objectives (*7_Objectives*), statistical methods for outcomes (*20a_Statistical_methods_Outcomes*), registry numbers (*3a_Registry_number*), and consent information (*26a_Consent_Obtaining*), all reported in more than 97% of the articles. The least frequently reported items were related to consent provisions (*26b_Consent_Provisions*), code sharing (*31e_Sharing_Code*), and consent materials (*32_Informed_Consent_Materials*) reported in fewer than 4% of relevant articles.

### Sentence-level dataset

The sentence-level dataset contains 52,294 sentences, including 6,777 section headers. 26,613 sentences (58.5%, excluding section headers) were annotated with checklist items. Each annotated sentence was annotated with an average of 1.16 items. The average number of annotations per sentence, including those with no labels, was 0.68. Item-specific statistics for the sentence-level dataset are also provided in Supplementary Table S2. Some items were reported over many sentences in a paper (e.g., *11a_Intervention_Description*, 20.36 sentences on average; *12a_Outcomes_Definitions*, 19.15 sentences); however, most items include at most a few sentences per article (e.g., *16a_Randomization_Generation, 16c_Randomization_Block_size*).

### Term-level dataset

The term-level dataset includes a total of 4,231 annotations, for an average of 21.16 terms per article. Out of 22 items, on average, 13.27 (±3.05) were annotated per article. In more than 90% of the articles, we annotated registry number (*3a_Registry_Number*), population/intervention in the title (*1e_Title_Population* and *1f_Title_Intervention*), and sample size (*14a_Sample_Size*). The least frequent terms were title-related: framework (*1c_Title_Framework*) and centers (*1d_Title_Centers*) are reported in less than 10% of the titles. Terms related to masked people (*17a_Masking_People_Masked*) were most frequently annotated (381 instances).

Terms related to statistical methods (*20c_Statistical_Methods_Analysis_Population, 20d_Statistical_Methods_Missing_Data*) include a noticeably larger number of tokens per term (up to 72 tokens for the former and 45 for the latter). 128 annotations (3%) have disjoint spans (e.g., in the title “Enteral vs. intravenous ICU sedation management*”, “*Enteral … ICU sedation management” is annotated with the item *1f_Title_Intervention*). Detailed descriptive statistics of the term-level dataset are provided in Supplementary Table S3.

### Technical Validation

In this section, we validate the corpus by reporting the inter-annotator agreement on articles annotated in duplicate. We also benchmark baseline NLP model performance using both established and novel metrics.

### Inter-annotator agreement (IAA)

We calculated IAA for annotations at the article, sentence, and term levels across three stages of annotation (5, 14, and 11 trials, respectively). At the article and sentence levels, IAA was calculated using Krippendorff’s α ^40^, which accommodates binary (article) and multi-label (sentence) cases. Simple binary distance was used at the article level, while MASI metric^41^ (incorporating Jaccard distance) was used at the sentence level to account for set overlap. As shown in Table 2, IAA improved across stages: from 0.682 to 0.773 at the article level and from 0.566 to 0.662 at the sentence level. The latter is higher than agreement reported in prior work on CONSORT checklist items^14,^ supporting subsequent single annotation.

**Table 2.**
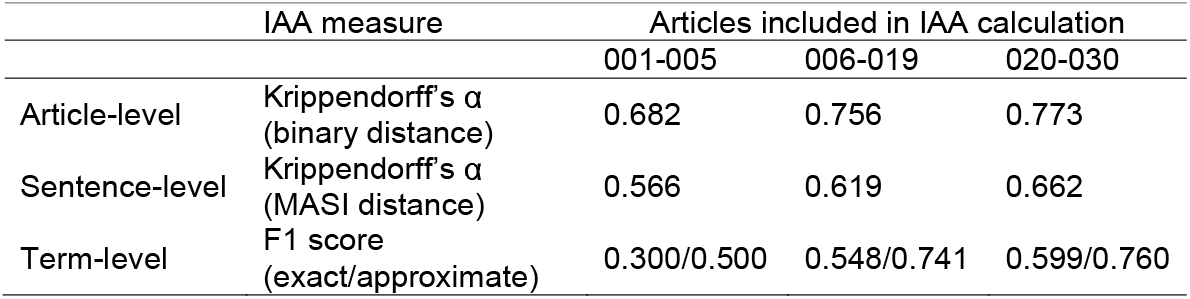
IAA calculated at different levels of the corpus. 30 trials (60 publications) are included in IAA calculation.

For the term-level dataset, we calculated IAA using F1 score^42,^ treating one annotator’s labels as ground truth and the other’s as predictions. Both exact and approximate matching (allowing term overlaps) were considered. In stage 1 annotation (articles 001-005), IAA averaged 0.3 (exact) and 0.5 (approximate). In stage 2 (articles 006-019), IAA improved to 0.548 (exact) and 0.741 (approximate). In stage 3 (articles 020-030), IAA further increased to 0.599 (exact) and 0.76 (approximate).

### NLP models and evaluation

We used the annotated sentence-level and term-level datasets to train NLP models that predict the reporting of checklist items. In this subsection, we describe the NLP methods used, evaluation metrics, and report performance of the models. We did not train a separate article-level model; article-level binary predictions were simply derived from sentence-level predictions.

#### Sentence-level prediction model

For sentence-level predictions, we retrained the multi-label text classification model that yielded best performance in our prior work^34^. The model encodes the input text using the PubMedBERT pre-trained encoder^43^ and feeds the resulting [CLS] token representation into a sigmoid-activated classification head for final prediction. The input text consists of three sentences (preceding, target, and trailing sentences) separated by [SEP] tokens and prepended by the [CLS] token. The corresponding section headers are also prepended to the start of each sentence. We refer the reader to Jiang et al.^34^ for further details on the model. To make the most efficient use of the annotated data, we trained a single model using 82 items. For the remaining item (*2_Abstract_structured*), which is an article-level item only and indicates whether the article includes a structured abstract, we integrated a rule-based method developed in previous work^34^. Despite developing a single comprehensive model, our evaluation considers SPIRIT and CONSORT subsets of the checklist items on protocol and results publications, respectively. This is because a user of this model is most likely to assess a protocol using SPIRIT items or a final report using CONSORT items, rather than using all 83 items for assessing adherence. For the experiments, we trained and evaluated the model 5 times on a NVIDIA V100-32GB GPU. We set the number of epochs to 20 and used a batch size of 4 for each run following prior work^34^. We report the evaluation metrics as mean average of 5 runs and provide 95% confidence intervals based on bootstrap sampling.

To account for different potential use cases for the sentence-level models, we used standard text classification evaluation metrics (precision, recall, and F1, both micro- and macro-averaged) across sentence and article levels. Sentence-level performance was calculated following the standard procedure for multi-label sentence classification tasks. Article-level performance was calculated by assessing whether, for a given checklist item, the label of *at least one* sentence is predicted correctly within the article. This evaluation metric is more lenient than sentence-level evaluation, although it facilitates practical use cases such as reporting checks and large-scale reporting analyses^35^, where the user would primarily focus on which checklist items are reported or missing and what evidence the model provides for the prediction.

The model performance at the sentence and article levels for all items as well as for SPIRIT and CONSORT items specifically is presented in Table 3. Sentence-level performance on the CONSORT checklist is higher than performance reported in prior work^34^; micro-F1 (0.748 vs. 0.71) and macro-F1 (0.701 vs. 0.67). The performance on the SPIRIT checklist is similar to that on CONSORT in terms of micro-F1 (0.748) but is lower in macro-F1 (0.668). At the article level, the model performs better on CONSORT than on SPIRIT for both micro-F1 (0.921 vs. 0.894) and macro-F1 (0.858 vs. 0.810). For CONSORT, article-level performance is also higher compared to that reported in prior work (0.90 micro-F1 and 0.84 macro-F1)^34^. Item-level results for the all-items model at the sentence and article levels are presented in Supplementary Tables S4-5, respectively. Analyzing the model predictions, we observe that the model does not perform well on infrequently reported items (e.g., *30_Post_trial_care*), consistent with our prior work^14,34^, and often confuses labels that are similar (e.g., *10a_Participants_inclusion* and *10b_Center_interventionist_inclusion*). While the performance, especially at the article level, seems reasonable for practical use, there is room for improving the model for the sentence-level predictions.

**Table 3.** Performance of the sentence classification model at sentence and article levels. Macro- and micro-averaged performance is reported, with 95% CIs in square brackets. SPIRIT-only performance is calculated by restricting the model predictions to protocols and to the items included in SPIRIT. Similarly, CONSORT-only performance relates to results publications and items included in CONSORT. All-items performance considers all 83 items and all publications.

|  |  | Precision [95% CI] | Recall [95% CI] | F1 [95% CI] |
| --- | --- | --- | --- | --- |
| *Sentence-level* |  |  |  |  |
| All-items | Micro | 0.756 [0.748-0.763] | 0.729 [0.723-0.737] | 0.742 [0.741-0.744] |
|  | Macro | 0.702 [0.696-0.708] | 0.624 [0.622-0.627] | 0.645 [0.641-0.648] |
| SPIRIT-only | Micro | 0.759 [0.748-0.769] | 0.738 [0.733-0.746] | 0.748 [0.745-0.751] |
|  | Macro | 0.712 [0.706-0.719] | 0.661 [0.654-0.668] | 0.668 [0.663-0.672] |
| CONSORT-only | Micro | 0.767 [0.758-0.774] | 0.729 [0.720-0.740] | 0.748 [0.744-0.751] |
|  | Macro | 0.743 [0.721-0.762] | 0.683 [0.672-0.693] | 0.701 [0.692-0.708] |
| *Article-level* |  |  |  |  |
| All-items | Micro | 0.887 [0.880-0.892] | 0.845 [0.842-0.847] | 0.865 [0.862-0.867] |
|  | Macro | 0.799 [0.795-0.803] | 0.744 [0.741-0.748] | 0.761 [0.757-0.764] |
| SPIRIT-only | Micro | 0.917 [0.910-0.923] | 0.871 [0.864-0.876] | 0.894 [0.890-0.898] |
|  | Macro | 0.846 [0.842-0.853] | 0.793 [0.784-0.802] | 0.810 [0.806-0.814] |
| CONSORT-only | Micro | 0.924 [0.917-0.931] | 0.918 [0.914-0.923] | 0.921 [0.916-0.925] |
|  | Macro | 0.875 [0.854-0.892] | 0.859 [0.853-0.865] | 0.858 [0.847-0.865] |

#### Term-level prediction model

We formulated term-level information extraction as a named entity recognition (NER) task. Specifically, we used a span prediction approach to NER, where the input consists of consecutive tokens from a sentence up to a fixed length *L* (i.e., candidate terms) to be classified into a term-level label (or None). For span prediction, we fine-tuned the PURE model^44^ on our term-level dataset. In the PURE model, a candidate term of length *k* (Tk = {*x*_*1*_,*x*_*2*_, …,*x*_*k*_}), where *x*_*i*_ is the *i*-th token of the term and *k ≤ L*, is represented as the concatenation of the contextualized representations of the first and the last tokens of the span as well as the trained embedding for the span length: *e*_*k*_ = [h_x1_ ;h_xk_;*φ(T*_*k*_)]. Here, *h*_*x*_ is the representation of the token *x* and *φ(T*_*k*_) corresponds to the learned embeddings of span width k. This span representation is then fed into a feedforward network to predict terms. We use PubMedBERT^43^ to generate contextualized token representations. We prefer span prediction approach to the more common BIO (Beginning-Inside-Outside) representation and token classification, as the terms in our corpus tend to be long phrases, unlike typical named entities, and the corpus included a considerable number of nested entities, which can be more naturally handled using span prediction. We excluded term mentions with disjoint spans from training and evaluation, as they are incompatible with a span prediction formulation for NER.

In addition to applying the baseline PURE model, we also examined whether the section headers and relative positions of the spans could improve the performance, as they can provide clues to the presence of specific terms. After some initial experiments to find which section headers to use, we prepended top-level section header to the input sentence (e.g. “Section-header: Abstract, Sentence: Results from previous studies on acupuncture for labour pain are contradictory and lack important information on methodology”). To represent relative positions of sentences, we first segmented each document into *k* chunks and assigned the relative position index to every sentence. We then encoded the sentence-level index of *T*_*k*_ as one-hot encoding as input to a feedforward network *ψ(T*_*k*_) to generate representation for the relation position and added this representation to the end of the span embeddings: *e*_*k*_ = [h_*x1*_ ;h_*xk*_;*φ(T*_*k*_);*ψ(T*_*k*_)]. Span prediction approach requires sampling of negative examples. We included all negative samples from sentences in titles, abstracts, and methods sections. In another experiment, we also sampled instances from sentences with positive labels only. In practice, we envision that a term-level model would be applied after sentence classification; therefore, using positive sentences only could be considered an upper bound for performance.

We used the validation set for hyperparameter tuning. We fixed maximum span length *L* to 10 tokens, as this covered about 95% of the term annotations. Unlike PURE, we did not include preceding and trailing sentences of the target sentence in training. We set the number of epochs as 200 and stopped training when F1 score did not improve compared to the previous 5 epochs. We used 4 NVIDIA V100-32GB GPUs with a batch size of 32. For the rest of the hyperparameters (e.g., learning rate), we followed the original hyperparameters of the PURE model.

To measure term extraction performance, we use standard NER metrics: precision, recall, and F1 score. We compute evaluation metrics in both strict vs. lenient modes and use positive sentences vs. all sentences as input. In strict evaluation, only exact match of the predicted span and ground truth span along with the term type match is considered correct, whereas lenient evaluation allows span overlaps but also requires term type match. We put more emphasis on lenient evaluation, because some term-level items tend to be expressed in long phrases (e.g., for *1e_Title_Population*: “patients undergoing coronary artery stenting for an acute coronary syndrome”) and overlap of spans could be considered acceptable in such cases.

Table 4 shows the high-level performance of our baseline term extraction models. The baseline PURE model yields 0.462 F1 score in strict evaluation when all test sentences are used and 0.505 F1 score when only positive sentences are used. The performance is 9-10 absolute percentage points higher in lenient evaluation (0.553 and 0.604, respectively), indicating that capturing term boundaries accurately is a significant challenge for the model. Precision increases significantly in positive sentence only evaluation compared to all sentences evaluation, indicating that sentence-level classification before applying the term extraction model would improve performance. Prepending section headers to inputs and adding relative position information improves the results over the baseline PURE model by 8.3-11 absolute percentage points, with a significant improvement in recall with some drop in precision. Overall, models perform better in precision compared to recall and further recall improvements would pave the way for practical use of the model. Item-level results are presented in Supplementary Table S6.

**Table 4.** Performance of the term extraction models. Mean averages over 5 runs with different seeds are shown, along with 95% CIs in square brackets. Note that term mentions with disjoint spans were excluded from evaluation.

| Model | Strict |  |  | Lenient |  |  |
| --- | --- | --- | --- | --- | --- | --- |
|  | Precision | Recall | F1 | Precision | Recall | F1 |
| *With samples from all sentences* |  |  |  |  |  |  |
| Baseline (PURE) | 0.572<br>[0.550-0.589] | 0.390<br>[0.356-0.432] | 0.462<br>[0.441-0.489] | 0.692<br>[0.676-0.707] | 0.463<br>[0.421-0.511] | 0.553<br>[0.524-0.584] |
| Our model | 0.554<br>[0.544-0.570] | 0.537<br>[0.530-0.542] | 0.545<br>[0.539-0.552] | 0.683<br>[0.669-0.699] | 0.644<br>[0.637-0.650] | 0.663<br>[0.655-0.669] |
| *With samples from sentences including term annotations only* |  |  |  |  |  |  |
| Baseline (PURE) | 0.722<br>[0.697-0.745] | 0.390<br>[0.356-0.432] | 0.505<br>[0.479-0.540] | 0.878<br>[0.861-0.892] | 0.463<br>[0.421-0.511] | 0.604<br>[0.569-0.643] |
| Our model | 0.714<br>[0.699-0.728] | 0.537<br>[0.531-0.542] | 0.613<br>[0.604-0.622] | 0.886<br>[0.881-0.889] | 0.644<br>[0.637-0.650] | 0.746<br>[0.741-0.750] |

Items such as *1a_Title_Randomized, 1g_Title_Acronym, 3a_Registry_Number*, and *17c_Masking_Type* perform well in both strict and lenient evaluations due to their more standardized mentions, which share similar linguistic and contextual patterns. In contrast, items like *1e_Title_Population, 1f_Title_Intervention*, and *16a_Randomization_Generation* show larger discrepancies between strict and lenient scores, as their mentions involve longer tokens (Supplementary Table S3), making exact boundaries harder to determine. F1 scores are lower for items such as *8b_Design_Framework, 17b_Masking_Not_masked, 20c_Statistical_methods_Analysis_population*, and *20d_Statistical_methods_Missing_data*. For *20c_Statistical_methods_Analysis_population* and *20d_Statistical_methods_Missing_data*, this seems partly due to their extremely long mentions. However, *8b_Design_Framework* and *17b_Masking_Not_masked* lag despite having ample training instances and moderate mention lengths.

Misclassification of design frameworks stems from comparative terms (e.g., ‘greater,’ ‘better’), which are often incorrectly identified across the text, leading to low precision. Similarly, *17b_Masking_Not_masked* is often misclassified as *17a_Masking_People_masked*, likely due to terms like ‘participants’ and ‘patients’, common for both items. While sentence-level prediction leverages context to distinguish these labels, term extraction relies more heavily on token-level features (e.g., start/end tokens and relative positions), which lack contextual depth. First filtering sentences predicted to contain items and then applying term extraction, could improve performance.

### Strengths and Limitations

We curated an expert-annotated corpus of 200 publications (100 trials) and more than 26K sentences with 83 checklist items related to SPIRIT and CONSORT reporting guidelines, making SPIRIT-CONSORT-TM the largest and most fine-grained publicly available corpus of its kind. Our team annotated part of the corpus iteratively and in duplicate to ensure consistency and quality in the corpus. We split the corpus into training, validation, and test splits and trained NLP models, ensuring that the corpus can serve as a benchmark and the models as baseline models for transparency assessment according to SPIRIT and CONSORT guidelines. Our baseline models show reasonable performance, although there is room for improvement.

Our corpus also has some limitations. All included publications were available in PubMed Central, which may not be representative of all RCT publications, although we aimed to include RCT publications on a broad range of topics. Some checklist items are reported infrequently and thus not well-represented in the corpus; NLP models can be expected to underperform on such labels. While the corpus is appropriate for identifying text related to SPIRIT or CONSORT items, we did not annotate whether each item was reported as recommended in the guidelines. We focused on parallel group trials about intervention effectiveness because those are the trials to which SPIRIT and CONSORT apply directly; future work might consider other types of trials for which SPIRIT and CONSORT extensions are available. Finally, the NLP models were only evaluated on our curated test set, and further external validation is needed to assess their generalizability. We are currently developing a web-based tool that will allow authors, journal staff, and others to upload manuscripts and publications and provide a report on reporting transparency based on the models. This, in addition to ongoing work on improving the model performance, will allow us to conduct robust external validation.

## Data Availability

Code, data, and materials related to the searching and processing of PubMed search results and screening of articles are available from https://osf.io/8rg4h/. Code used for training and evaluating the models is available at https://github.com/ScienceNLP-Lab/RCT-Transparency/tree/main/SPIRIT-CONSORT-TM.

## Code Availability

Code, data, and materials related to the searching and processing of PubMed search results and screening of articles are available from https://osf.io/8rg4h/. Code used for training and evaluating the models is available at https://github.com/ScienceNLP-Lab/RCT-Transparency/tree/main/SPIRIT-CONSORT-TM. This repository contains an environment file specifying the versions of any software used during this process, as well as a configuration file containing the parameters used in the experiments.

## Acknowledgements

This work was supported by the National Library of Medicine of the National Institutes of Health under the award number R01LM014079. The content is solely the responsibility of the authors and does not necessarily represent the official views of the National Institutes of Health. The funder had no role in considering the study design or in the collection, analysis, interpretation of data, writing of the report, or decision to submit the article for publication. This work used Bridges-2 and Ocean at Pittsburgh Supercomputing Center (PSC) through allocation CIS230380 from the Advanced Cyberinfrastructure Coordination Ecosystem: Services Support (ACCESS) program^45^, which is supported by National Science Foundation, United States grants #2138259, #2138286, #2138307, #2137603, and #2138296.

## Author contributions

LJ: Methodology, Data curation, Software, Validation, Formal analysis, Investigation, Writing – Original draft, Writing – Review & Editing. CJV: Conceptualization, Methodology, Data curation, Writing – Original draft, Writing – Review & Editing. XY: Conceptualization, Methodology, Data curation, Writing – Review & Editing. AWB: Conceptualization, Methodology, Data curation, Writing – Review & Editing. JDM: Methodology, Data curation, Software, Validation, Formal analysis, Investigation, Writing – Original draft, Writing – Review & Editing. GH: Methodology, Data curation, Software, Validation, Formal analysis, Investigation, Writing – Original draft, Writing – Review & Editing. ML: Methodology, Software, Validation, Writing – Original draft, Writing – Review & Editing. EM-W: Conceptualization, Methodology, Data curation, Supervision, Project administration, Funding acquisition, Writing – Review & Editing. HK: Conceptualization, Methodology, Investigation, Supervision, Project administration, Funding acquisition, Writing – Original draft, Writing – Review & Editing.

