## Supplementary Table for "SPIRIT-CONSORT-TM: a corpus for assessing transparency of clinical trial protocol and results publications"

We provide token-level statistics for the training, validation, and test splits of the SPIRIT-CONSORT-TM corpus, along with item-level details, and item-level performance of the NLP models below.

Table S1 presents the token-level statistics of the dataset across training, validation, and test splits.

| Split | Total no. of tokens | Mean (SD) | Median (IQR) |
|---|---|---|---|
| Training | 854,289 | 6,102.06 (1,953.11) | 5,918.00 (2,003.50) |
| Validation | 108,870 | 5,443.50 (1,094.30) | 5,695.00 (1,434.25) |
| Test | 247,948 | 6,198.70 (1,870.28) | 6,098.00 (1,621.50) |

**Table S1**. Token-level descriptive statistics of SPIRIT-CONSORT-TM. SD: standard deviation.

Table S2 presents descriptive statistics of the dataset at the sentence and article levels.

| Checklist Item | No. of articles | Avg. number of sentences per article (SD) | Range |
|---|---|---|---|
| 1a_Title_Randomized | 168 | 0.85 (0.39) | 0-2 |
| 1b_Title_Type | 33 | 0.17 (0.37) | 0-1 |
| 1c_Title_Framework | 17 | 0.09 (0.28) | 0-1 |
| 1d_Title_Centers | 19 | 0.10 (0.29) | 0-1 |
| 1e_Title_Population | 188 | 0.95 (0.26) | 0-2 |
| 1f_Title_Intervention | 189 | 0.95 (0.24) | 0-2 |
| 1g_Title_Acronym | 68 | 0.34 (0.47) | 0-1 |
| 2_Abstract_structured | 184 | 0.92 (0.27) | 0-1 |
| 3a_Registry_number | 195 | 1.44 (0.68) | 0-5 |
| 3b_Protocol_access** | 90 | 0.81 (1.15) | 0-6 |
| 4_Funding | 188 | 2.71 (2.54) | 0-20 |
| 5a_Sponsor | 30 | 0.21 (0.55) | 0-3 |
| 5b_Contributors_roles | 153 | 5.22 (4.59) | 0-36 |
| 5c_Oversight_committees | 43 | 1.52 (9.03) | 0-87 |
| 7_Objectives | 198 | 2.85 (2.48) | 0-13 |
| 8a_Design_Type | 105 | 0.70 (0.83) | 0-5 |
| 8b_Design_Framework | 134 | 1.12 (1.15) | 0-6 |
| 8c_Design_Centers | 177 | 1.35 (0.87) | 0-4 |
| 8d_Design_Ratio | 148 | 0.99 (0.79) | 0-4 |
| 9_Setting | 186 | 2.22 (3.33) | 0-31 |
| 10a_Participants_inclusion | 200 | 5.70 (6.18) | 1-40 |
| 10b_Center_interventionist_inclusion | 50 | 0.58 (1.56) | 0-11 |
| 11a_Intervention_Description | 199 | 20.36 (19.74) | 0-122 |
| 11b_Intervention_Modification | 45 | 0.50 (1.44) | 0-14 |
| 11c_Intervention_Monitoring | 124 | 2.62 (4.97) | 0-45 |
| 11d_Intervention_Concomitant | 104 | 1.63 (3.78) | 0-36 |

| | | | |
|---|---|---|---|
| 12a_Outcomes_Definitions | 198 | 19.15 (19.42) | 0-116 |
| 12b_Outcomes_Changes** | 11 | 0.09 (0.44) | 0-4 |
| 13_Participant_timeline | 103 | 1.85 (5.33) | 0-60 |
| 14a_Sample_size | 183 | 1.42 (1.13) | 0-11 |
| 14b_Sample_Calculation | 186 | 5.44 (6.35) | 0-44 |
| 15_Recruitment | 143 | 2.51 (3.51) | 0-26 |
| 16a_Randomization_Generation | 143 | 0.78 (0.54) | 0-2 |
| 16b_Randomization_Type | 142 | 0.99 (0.87) | 0-6 |
| 16c_Randomization_Block_size | 73 | 0.40 (0.56) | 0-2 |
| 16d_Randomization_Strata | 104 | 0.62 (0.68) | 0-3 |
| 16e_Allocation_Mechanism | 127 | 0.76 (0.89) | 0-10 |
| 16f_Allocation_Concealment | 133 | 1.10 (1.22) | 0-9 |
| 16g_Personnel_Sequence | 119 | 0.70 (0.72) | 0-5 |
| 16h_Personnel_Enrollment | 96 | 0.74 (1.00) | 0-6 |
| 17a_Masking_People_masked | 134 | 1.20 (1.23) | 0-7 |
| 17b_Masking_Not_masked | 93 | 0.56 (0.68) | 0-3 |
| 17c_Masking_Type | 109 | 0.78 (0.93) | 0-5 |
| 17d_Masking_Unblinding | 22 | 0.21 (0.96) | 0-9 |
| 17e_Masking_Similarity | 55 | 0.49 (0.99) | 0-5 |
| 18a_Data_Collection | 174 | 7.97 (11.44) | 0-84 |
| 18b_Data_Retention | 76 | 0.87 (1.55) | 0-9 |
| 19_Data_Management | 79 | 1.43 (3.13) | 0-23 |
| 20a_Statistical_methods_Outcomes | 196 | 7.71 (7.43) | 0-48 |
| 20b_Statistical_methods_Other_Analyses | 101 | 1.52 (2.79) | 0-19 |
| 20c_Statistical_methods_Analysis_population | 143 | 1.35 (1.35) | 0-8 |
| 20d_Statistical_methods_Missing_data | 89 | 0.74 (1.12) | 0-7 |
| 21a_Data_monitoring_committee | 58 | 0.88 (2.26) | 0-20 |
| 21b_Interim_analyses | 42 | 0.52 (1.79) | 0-19 |
| 21c_Stopping_guidelines | 28 | 0.40 (2.38) | 0-32 |
| 22_Harms_non-systematic | 85 | 1.55 (4.16) | 0-39 |
| 23_Auditing* | 18 | 0.27 (1.01) | 0-7 |
| 24_Ethics | 193 | 1.83 (2.48) | 0-32 |
| 25_Amendments | 47 | 0.53 (1.44) | 0-14 |
| 26a_Consent_Obtaining | 194 | 2.40 (2.89) | 0-27 |
| 26b_Consent_Provisions* | 4 | 0.04 (0.27) | 0-3 |
| 27_Confidentiality | 72 | 0.85 (1.83) | 0-14 |
| 28_Financial_interests | 170 | 2.62 (4.46) | 0-46 |
| 29_Data_access* | 24 | 0.19 (0.60) | 0-4 |
| 30_Post_trial_care* | 16 | 0.17 (0.77) | 0-8 |
| 31a_Dissemination | 37 | 0.57 (1.54) | 0-11 |
| 31b_Authorship | 14 | 0.12 (0.59) | 0-7 |
| 31c_Sharing_Materials | 24 | 0.23 (0.71) | 0-6 |
| 31d_Sharing_Data | 82 | 0.99 (1.55) | 0-13 |
| 31e_Sharing_Code | 7 | 0.05 (0.30) | 0-3 |
| 32_Informed_consent_materials | 8 | 0.05 (0.28) | 0-3 |
| 33_Biological_specimens | 14 | 0.29 (1.30) | 0-10 |
| 34_Flow** | 98 | 2.16 (2.74) | 0-12 |
| 35a_Recruitment_dates | 145 | 0.95 (0.78) | 0-4 |
| 35b_Followup_dates | 74 | 0.43 (0.61) | 0-3 |
| 35c_Stopping** | 15 | 0.11 (0.49) | 0-5 |
| 36_Baseline_data** | 100 | 2.46 (3.65) | 0-16 |
| 37a_Analysis_Numbers** | 82 | 0.79 (1.33) | 0-10 |
| 38a_Outcome_results** | 99 | 9.20 (13.11) | 0-70 |

| | | | |
|---|---|---|---|
| 38b_Binary_results** | 64 | 1.55 (4.11) | 0-44 |
| 39_Ancillary_results** | 76 | 3.31 (8.01) | 0-88 |
| 40_Harms_results** | 47 | 1.06 (2.87) | 0-23 |
| 41_Generalizability** | 81 | 1.07 (1.95) | 0-12 |

**Table S2**. Sentence-level descriptive statistics of checklist items in SPIRIT-CONSORT-TM. SD: standard deviation. *: SPIRIT-only items, **: CONSORT-only items.

Table S3 shows the descriptive statistics of the term-level dataset at the item level.

| Checklist Item | No. of articles | No. of instances | Avg. frequency of terms per article (SD) | Range (# of annotations) | Avg. length of tokens (SD) | Range (token length) |
|---|---|---|---|---|---|---|
| 1a_Title_Randomized | 168 | 172 | 0.86 (0.40) | 0-2 | 1.02 (0.23) | 1-4 |
| 1b_Title_Type | 33 | 34 | 0.17 (0.39) | 0-2 | 1.47 (0.85) | 1-3 |
| 1c_Title_Framework | 17 | 20 | 0.10(0.35) | 0-2 | 1.15 (0.36) | 1-2 |
| 1d_Title_Centers | 19 | 19 | 0.10 (0.29) | 0-1 | 1.74 (0.91) | 1-3 |
| 1e_Title_Population | 188 | 210 | 1.05 (0.48) | 0-4 | 4.30 (3.06) | 1-18 |
| 1f_Title_Intervention | 189 | 245 | 1.23 (0.64) | 0-4 | 4.24 (2.99) | 1-16 |
| 1g_Title_Acronym | 68 | 68 | 0.34 (0.47) | 0-1 | 1.71 (1.09) | 1-7 |
| 3a_Registry_number | 195 | 304 | 1.52 (0.82) | 0-6 | 1.19 (1.00) | 1-13 |
| 8a_Design_Type | 105 | 153 | 0.77 (0.99) | 0-7 | 2.24 (1.77) | 1-11 |
| 8b_Design_Framework | 134 | 261 | 1.30 (1.54) | 0-10 | 2.94 (4.85) | 1-31 |
| 8c_Design_Centers | 177 | 279 | 1.40 (0.96) | 0-6 | 2.75 (2.01) | 1-17 |
| 8d_Design_Ratio | 148 | 206 | 1.03 (0.87) | 0-5 | 4.00 (2.10) | 2-14 |
| 14a_Sample_size | 183 | 357 | 1.78 (1.42) | 0-13 | 3.05 (3.24) | 1-29 |
| 16a_Randomization_Generation | 143 | 154 | 0.77 (0.54) | 0-2 | 4.55 (4.01) | 1-22 |
| 16b_Randomization_Type | 142 | 240 | 1.20 (1.06) | 0-6 | 1.49 (1.29) | 1-12 |
| 16c_Randomization_Block_size | 73 | 84 | 0.42 (0.60) | 0-3 | 3.48 (3.15) | 1-12 |
| 16d_Randomization_Strata | 104 | 218 | 1.09 (1.39) | 0-8 | 6.07 (7.16) | 1-43 |
| 17a_Masking_People_masked | 134 | 381 | 1.91 (2.04) | 0-13 | 2.15 (2.10) | 1-21 |
| 17b_Masking_Not_masked | 93 | 167 | 0.83 (1.09) | 0-6 | 2.08 (1.84) | 1-10 |
| 17c_Masking_Type | 109 | 157 | 0.79 (0.94) | 0-5 | 2.74 (0.88) | 1-9 |
| 20c_Statistical_methods_Analysis_population | 143 | 352 | 1.76 (2.01) | 0-16 | 7.28 (7.97) | 1-72 |
| 20d_Statistical_methods_Missing_data | 89 | 150 | 0.75 (1.14) | 0-7 | 7.89 (8.26) | 1-45 |

**Table S3.** Descriptive statistics regarding the annotation of term-level checklist items in SPIRIT-CONSORT-TM. SD: standard deviation.

Tables S4 and S5 show the NLP model performance over 5 runs at the sentence and article levels. 95% CIs are not included for brevity.

| Checklist Item | Prec. | Recall | F1 |
|---|---|---|---|
| 1a_Title_Randomized | 0.899 | 1.000 | 0.946 |
| 1b_Title_Type | 1.000 | 0.500 | 0.667 |

| | | | |
|---|---|---|---|
| 1c_Title_Framework | 0.000 | 0.000 | 0.000 |
| 1d_Title_Centers | 1.000 | 0.933 | 0.960 |
| 1e_Title_Population | 0.897 | 0.979 | 0.936 |
| 1f_Title_Intervention | 0.917 | 0.959 | 0.937 |
| 1g_Title_Acronym | 0.986 | 0.943 | 0.964 |
| 3a_Registry_number | 0.900 | 0.949 | 0.923 |
| 3b_Protocol_access | 0.824 | 0.766 | 0.793 |
| 4_Funding | 0.894 | 0.906 | 0.900 |
| 5a_Sponsor | 0.767 | 0.680 | 0.708 |
| 5b_Contributors_roles | 0.984 | 0.963 | 0.973 |
| 5c_Oversight_committees | 0.584 | 0.700 | 0.635 |
| 7_Objectives | 0.835 | 0.860 | 0.845 |
| 8a_Design_Type | 0.929 | 0.439 | 0.593 |
| 8b_Design_Framework | 0.598 | 0.621 | 0.608 |
| 8c_Design_Centers | 0.633 | 0.512 | 0.566 |
| 8d_Design_Ratio | 0.822 | 0.657 | 0.730 |
| 9_Setting | 0.801 | 0.611 | 0.693 |
| 10a_Participants_inclusion | 0.908 | 0.874 | 0.891 |
| 10b_Center_interventionist_inclusion | 0.500 | 0.227 | 0.303 |
| 11a_Intervention_Description | 0.807 | 0.910 | 0.855 |
| 11b_Intervention_Modification | 0.240 | 0.100 | 0.133 |
| 11c_Intervention_Monitoring | 0.591 | 0.543 | 0.561 |
| 11d_Intervention_Concomitant | 0.687 | 0.337 | 0.451 |
| 12a_Outcomes_Definitions | 0.787 | 0.672 | 0.724 |
| 12b_Outcomes_Changes** | 0.000 | 0.000 | 0.000 |
| 13_Participant_timeline | 0.447 | 0.530 | 0.481 |
| 14a_Sample_size | 0.732 | 0.646 | 0.685 |
| 14b_Sample_Calculation | 0.876 | 0.855 | 0.865 |
| 15_Recruitment | 0.554 | 0.737 | 0.632 |
| 16a_Randomization_Generation | 0.881 | 0.893 | 0.887 |
| 16b_Randomization_Type | 0.818 | 0.844 | 0.828 |
| 16c_Randomization_Block_size | 0.969 | 0.871 | 0.917 |
| 16d_Randomization_Strata | 0.910 | 0.657 | 0.761 |
| 16e_Allocation_Mechanism | 0.581 | 0.539 | 0.556 |
| 16f_Allocation_Concealment | 0.587 | 0.594 | 0.588 |
| 16g_Personnel_Sequence | 0.642 | 0.627 | 0.632 |
| 16h_Personnel_Enrollment | 0.343 | 0.353 | 0.344 |
| 17a_Masking_People_masked | 0.857 | 0.816 | 0.836 |
| 17b_Masking_Not_masked | 0.797 | 0.520 | 0.626 |
| 17c_Masking_Type | 0.926 | 0.610 | 0.729 |
| 17d_Masking_Unblinding | 0.400 | 0.400 | 0.394 |
| 17e_Masking_Similarity | 0.661 | 0.346 | 0.453 |
| 18a_Data_Collection | 0.390 | 0.527 | 0.447 |
| 18b_Data_Retention | 0.845 | 0.473 | 0.606 |
| 19_Data_Management | 0.737 | 0.609 | 0.666 |
| 20a_Statistical_methods_Outcomes | 0.713 | 0.675 | 0.692 |
| 20b_Statistical_methods_Other_Analyses | 0.603 | 0.565 | 0.582 |
| 20c_Statistical_methods_Analysis_population | 0.783 | 0.694 | 0.735 |
| 20d_Statistical_methods_Missing_data | 0.785 | 0.639 | 0.703 |
| 21a_Data_monitoring_committee | 0.844 | 0.507 | 0.633 |
| 21b_Interim_analyses | 0.913 | 0.392 | 0.548 |
| 21c_Stopping_guidelines | 0.764 | 0.919 | 0.832 |
| 22_Harms_non-systematic | 0.863 | 0.836 | 0.848 |

| | | | |
|---|---|---|---|
| 23_Auditing* | 0.383 | 0.600 | 0.465 |
| 24_Ethics | 0.872 | 0.881 | 0.876 |
| 25_Amendments | 0.878 | 0.560 | 0.677 |
| 26a_Consent_Obtaining | 0.782 | 0.734 | 0.757 |
| 26b_Consent_Provisions* | N/A | N/A | N/A |
| 27_Confidentiality | 0.752 | 0.686 | 0.716 |
| 28_Financial_interests | 0.952 | 0.957 | 0.954 |
| 29_Data_access* | 0.755 | 0.767 | 0.744 |
| 30_Post_trial_care | 0.333 | 0.050 | 0.084 |
| 31a_Dissemination | 0.819 | 0.900 | 0.857 |
| 31b_Authorship | 0.828 | 0.533 | 0.640 |
| 31c_Sharing_Materials | 0.400 | 0.200 | 0.266 |
| 31d_Sharing_Data | 0.750 | 0.929 | 0.829 |
| 31e_Sharing_Code | N/A | N/A | N/A |
| 32_Informed_consent_materials | 0.000 | 0.000 | 0.000 |
| 33_Biological_specimens | 0.058 | 0.200 | 0.090 |
| 34_Flow** | 0.705 | 0.861 | 0.775 |
| 35a_Recruitment_dates | 0.818 | 0.842 | 0.830 |
| 35b_Followup_dates | 0.841 | 0.533 | 0.653 |
| 35c_Stopping | 0.800 | 0.266 | 0.400 |
| 36_Baseline_data | 0.816 | 0.683 | 0.741 |
| 37a_Analysis_Numbers | 0.306 | 0.261 | 0.271 |
| 38a_Outcome_results | 0.802 | 0.846 | 0.824 |
| 38b_Binary_results | 0.361 | 0.529 | 0.427 |
| 39_Ancillary_results | 0.760 | 0.481 | 0.588 |
| 40_Harms_results | 0.912 | 0.892 | 0.901 |
| 41_Generalizability | 0.469 | 0.433 | 0.450 |
| Micro-Average | 0.756 | 0.729 | 0.742 |
| Macro-Average | 0.702 | 0.624 | 0.645 |

**Table S4**. Item-level performance of the sentence classification model for each item. Items with NA for performance did not have any instances in the test set.

| Checklist Item | Prec. | Recall | F1 |
|---|---|---|---|
| 1a_Title_Randomized | 0.952 | 1.000 | 0.975 |
| 1b_Title_Type | 1.000 | 0.500 | 0.667 |
| 1c_Title_Framework | 0.000 | 0.000 | 0.000 |
| 1d_Title_Centers | 1.000 | 0.933 | 0.960 |
| 1e_Title_Population | 0.949 | 0.979 | 0.964 |
| 1f_Title_Intervention | 0.954 | 0.979 | 0.966 |
| 1g_Title_Acronym | 0.986 | 0.943 | 0.964 |
| 2_Abstract_structured | 0.971 | 0.971 | 0.971 |
| 3a_Registry_number | 0.974 | 0.995 | 0.984 |
| 3b_Protocol_access | 0.931 | 0.941 | 0.936 |
| 4_Funding | 0.979 | 0.984 | 0.982 |
| 5a_Sponsor | 0.767 | 0.760 | 0.737 |
| 5b_Contributors_roles | 1.000 | 0.933 | 0.966 |
| 5c_Oversight_committees | 0.762 | 0.785 | 0.769 |
| 7_Objectives | 0.979 | 0.944 | 0.961 |
| 8a_Design_Type | 0.948 | 0.615 | 0.743 |
| 8b_Design_Framework | 0.887 | 0.740 | 0.806 |
| 8c_Design_Centers | 0.856 | 0.662 | 0.745 |
| 8d_Design_Ratio | 0.930 | 0.810 | 0.865 |

| | | | |
|---|---|---|---|
| 9_Setting | 0.924 | 0.794 | 0.854 |
| 10a_Participants_inclusion | 1.000 | 0.975 | 0.987 |
| 10b_Center_interventionist_inclusion | 0.580 | 0.433 | 0.489 |
| 11a_Intervention_Description | 0.989 | 0.909 | 0.947 |
| 11b_Intervention_Modification | 0.327 | 0.249 | 0.279 |
| 11c_Intervention_Monitoring | 0.732 | 0.774 | 0.745 |
| 11d_Intervention_Concomitant | 0.660 | 0.609 | 0.631 |
| 12a_Outcomes_Definitions | 1.000 | 0.985 | 0.992 |
| 12b_Outcomes_Changes** | 0.000 | 0.000 | 0.000 |
| 13_Participant_timeline | 0.654 | 0.664 | 0.657 |
| 14a_Sample_size | 0.905 | 0.837 | 0.869 |
| 14b_Sample_Calculation | 0.967 | 1.000 | 0.983 |
| 15_Recruitment | 0.734 | 0.843 | 0.783 |
| 16a_Randomization_Generation | 0.963 | 0.911 | 0.936 |
| 16b_Randomization_Type | 0.929 | 0.918 | 0.922 |
| 16c_Randomization_Block_size | 0.969 | 0.938 | 0.953 |
| 16d_Randomization_Strata | 1.000 | 0.733 | 0.844 |
| 16e_Allocation_Mechanism | 0.705 | 0.597 | 0.642 |
| 16f_Allocation_Concealment | 0.816 | 0.745 | 0.776 |
| 16g_Personnel_Sequence | 0.686 | 0.793 | 0.735 |
| 16h_Personnel_Enrollment | 0.480 | 0.640 | 0.546 |
| 17a_Masking_People_masked | 1.000 | 0.924 | 0.960 |
| 17b_Masking_Not_masked | 0.931 | 0.564 | 0.703 |
| 17c_Masking_Type | 1.000 | 0.822 | 0.900 |
| 17d_Masking_Unblinding | 0.800 | 0.433 | 0.560 |
| 17e_Masking_Similarity | 0.749 | 0.531 | 0.619 |
| 18a_Data_Collection | 0.812 | 0.843 | 0.826 |
| 18b_Data_Retention | 0.880 | 0.715 | 0.785 |
| 19_Data_Management | 0.892 | 0.982 | 0.933 |
| 20a_Statistical_methods_Outcomes | 0.974 | 0.949 | 0.961 |
| 20b_Statistical_methods_Other_Analyses | 0.602 | 0.537 | 0.564 |
| 20c_Statistical_methods_Analysis_population | 0.902 | 0.842 | 0.869 |
| 20d_Statistical_methods_Missing_data | 0.924 | 0.708 | 0.800 |
| 21a_Data_monitoring_committee | 0.901 | 0.880 | 0.888 |
| 21b_Interim_analyses | 0.800 | 0.571 | 0.667 |
| 21c_Stopping_guidelines | 0.734 | 0.800 | 0.759 |
| 22_Harms_non-systematic | 0.875 | 0.789 | 0.829 |
| 23_Auditing* | 0.383 | 0.500 | 0.427 |
| 24_Ethics | 0.973 | 1.000 | 0.986 |
| 25_Amendments | 0.889 | 0.750 | 0.809 |
| 26a_Consent_Obtaining | 0.983 | 0.937 | 0.960 |
| 26b_Consent_Provisions* | N/A | N/A | N/A |
| 27_Confidentiality | 0.903 | 0.775 | 0.833 |
| 28_Financial_interests | 1.000 | 0.977 | 0.989 |
| 29_Data_access* | 0.750 | 0.800 | 0.755 |
| 30_Post_trial_care | 0.300 | 0.090 | 0.133 |
| 31a_Dissemination | 0.863 | 0.857 | 0.856 |
| 31b_Authorship | 0.767 | 0.600 | 0.662 |
| 31c_Sharing_Materials | 0.400 | 0.200 | 0.266 |
| 31d_Sharing_Data | 0.953 | 1.000 | 0.975 |
| 31e_Sharing_Code | N/A | N/A | N/A |
| 32_Informed_consent_materials | 0.000 | 0.000 | 0.000 |
| 33_Biological_specimens | 0.350 | 0.400 | 0.371 |

| | | | |
|---|---|---|---|
| 34_Flow** | 0.796 | 1.000 | 0.885 |
| 35a_Recruitment_dates | 0.881 | 0.926 | 0.902 |
| 35b_Followup_dates | 0.850 | 0.660 | 0.741 |
| 35c_Stopping | 0.800 | 0.266 | 0.400 |
| 36_Baseline_data | 0.918 | 1.000 | 0.957 |
| 37a_Analysis_Numbers | 0.400 | 0.435 | 0.406 |
| 38a_Outcome_results | 0.990 | 1.000 | 0.995 |
| 38b_Binary_results | 0.711 | 0.983 | 0.820 |
| 39_Ancillary_results | 0.695 | 0.759 | 0.725 |
| 40_Harms_results | 0.896 | 0.933 | 0.913 |
| 41_Generalizability | 0.636 | 0.710 | 0.671 |
| Micro-Average | 0.887 | 0.845 | 0.865 |
| Macro-Average | 0.799 | 0.744 | 0.761 |

**Table S5**. Article-level performance of the sentence classification model for each item. Items with NA for performance did not have any instances in the test set.

Table S6 shows the results of the term recognition model at the item level. 95% CIs are not included for brevity.

| **Checklist Item** | **Strict** | | | **Lenient** | | |
|---|---|---|---|---|---|---|
| | Prec. | Recall | F1 | Prec. | Recall | F1 |
| 1a_Title_Randomized | 1.000 | 1.000 | 1.000 | 1.000 | 1.000 | 1.000 |
| 1b_Title_Type | 1.000 | 0.500 | 0.667 | 1.000 | 0.500 | 0.667 |
| 1c_Title_Framework | 1.000 | 0.250 | 0.400 | 1.000 | 0.250 | 0.400 |
| 1d_Title_Centers | 1.000 | 0.333 | 0.500 | 1.000 | 0.333 | 0.500 |
| 1e_Title_Population | 0.514 | 0.475 | 0.494 | 0.892 | 0.825 | 0.857 |
| 1f_Title_Intervention | 0.568 | 0.532 | 0.550 | 0.857 | 0.766 | 0.809 |
| 1g_Title_Acronym | 0.846 | 0.786 | 0.815 | 0.923 | 0.857 | 0.889 |
| 3a_Registry_number | 0.873 | 0.965 | 0.917 | 0.889 | 0.983 | 0.933 |
| 8a_Design_Type | 0.611 | 0.393 | 0.478 | 0.706 | 0.429 | 0.533 |
| 8b_Design_Framework | 0.264 | 0.434 | 0.329 | 0.337 | 0.528 | 0.412 |
| 8c_Design_Centers | 0.568 | 0.438 | 0.494 | 0.595 | 0.458 | 0.518 |
| 8d_Design_Ratio | 0.774 | 0.585 | 0.667 | 0.903 | 0.683 | 0.778 |
| 14a_Sample_size | 0.521 | 0.576 | 0.547 | 0.548 | 0.606 | 0.576 |
| 16a_Randomization_Generation | 0.409 | 0.300 | 0.346 | 0.895 | 0.567 | 0.694 |
| 16b_Randomization_Type | 0.629 | 0.512 | 0.564 | 0.800 | 0.651 | 0.718 |
| 16c_Randomization_Block_size | 0.714 | 0.357 | 0.476 | 0.857 | 0.429 | 0.571 |
| 16d_Randomization_Strata | 0.500 | 0.303 | 0.377 | 0.737 | 0.424 | 0.538 |
| 17a_Masking_People_masked | 0.544 | 0.595 | 0.568 | 0.701 | 0.726 | 0.713 |
| 17b_Masking_Not_masked | 1.000 | 0.050 | 0.095 | 1.000 | 0.050 | 0.095 |
| 17c_Masking_Type | 0.838 | 0.775 | 0.805 | 0.865 | 0.800 | 0.831 |
| 20c_Statistical_methods_Analysis_population | 0.410 | 0.333 | 0.368 | 0.513 | 0.417 | 0.460 |
| 20d_Statistical_methods_Missing_data | 0.500 | 0.273 | 0.353 | 0.722 | 0.394 | 0.510 |
| OVERALL | 0.585 | 0.532 | 0.557 | 0.712 | 0.633 | 0.670 |

**Table S6**. Item-level performance from our term extraction model (PURE model with section header and relative position extension). Note that this table provides the item-level result from a single run; therefore the overall results differ from those in Table 4 of the main manuscript.
